# Exploring what lies beneath the tip of the gender-based violence iceberg

**DOI:** 10.1101/2024.02.26.24303373

**Authors:** David Moriña, Isabel Millán, Amanda Fernández-Fontelo, Pedro Puig, Pere Toran, Meritxell Gómez-Maldonado, Gemma Falguera

## Abstract

Gender-based violence refers to violence directed against a person because of that person’s gender or violence that affects persons of a particular gender disproportionately. It is estimated that 30% of women worldwide have suffered either physical and/or sexual violence in their lifetime. Primary Health Care could be one of the ideal places for the detection of these situations, but most of the cases remain undetected as the victims often decline to seek for medical care after suffering an event. This work shows that public primary health care system in Catalonia might be registering only around 50% of the cases currently, and it will take more than 20 years to see the whole picture of the phenomenon, and the situation could be the same in countries with similar socioeconomic contexts. We found in previous studies that gender-based violence cases are severely underregistered from the public health and judicial perspectives, on the basis of qualitative analyses and survey data. Furthermore, we propose a statistical modelling approach able to estimate the actual burden of this issue accurately. Our results show that awareness training campaigns focused on primary healthcare professionals are very effective in reducing the underreporting issue but should be conducted repeatedly and not only once.

## 1 Introduction

Gender-based violence (GBV), that is, violence directed against a person because of their gender or violence that affects persons of a particular gender disproportionately, can take various forms, including, most notoriously, domestic violence. Here, we are concerned specifically with physical or sexual violence directed against adult women, who are, jointly with girls, the main victims of GBV. According to the World Health Organization (WHO), 30% of women worldwide have suffered either physical and/or sexual violence at some point in their lives ([1]), and tackling this issue is one of the specific development goals of their 2030 agenda. It is clearly also a significant healthcare policy concern, as victims of sexual and physical GBV are more likely to require health services than the general population to address the physical, gynecological and psychological consequences of the aggressions suffered.

According to the macro survey on GBV violence conducted by the Spanish Government in 2019 ([2]), 21.5% of women in Spain aged 16 and older report having been victims of physical violence while 13.7% report having suffered acts of sexual violence. The findings of this survey stress both the ubiquity and severity of physical and sexual GBV in our society. However, it also reported that only 32% of the victims of physical and sexual GBV actually report the episode of violence to the police or justice system. This figure serves to highlight another facet to the problem that urgently needs to be addressed: The fact that most episodes of GBV go unreported or what has been referred to as the “GBV iceberg”.

Between 55 and 95% of women survivors of violence do not disclose or avail themselves of any type of health, legal or police services ([3]). In 2019, while it was estimated that 14.2% of women residing in Spain had suffered physical or sexual violence and that 21.7% of them had reported the situation to the police or judiciary, only 6.78% had gone to primary health services ([4]). Indeed, the latter should have a key role to play in the detection of GBV, given that women typically visit a primary healthcare unit at some point in their lives when seeking advice about sexual and reproductive health. Clearly, in this context, the work of healthcare professionals is vital ([5]). [6] seek to quantify the degree of underreporting of episodes of GBV –that is, as reported directly by the victim or friend/family member, by the social services (when they have pertinent information about the aggression) or by the police (when the victim files an official report)– for the provinces of Galicia (Spain) and find that underreporting is more severe in rural areas than it is in urban zones. The authors conclude that underreporting episodes of GBV undermines the quality of the associated data, resulting in poor, biased results, with the risk that society misunderstands the actual scope of the issue.

To address the GBV problem more effectively, we need to be able to identify all the cases of GBV that lie beneath the tip of the iceberg. To do so, one of the first questions we must address is why the victims of GBV fail to report episodes of violence. Two main reasons are typically forwarded: personal –i.e. the sense of shame, fear of retaliation, economic dependence, etc.– and societal –i.e. the imbalance in male/female power relations, protecting family privacy, victim blaming, etc. ([7]). As is evident, many of these reasons are closely associated with social stigma and form part of the victim’s religious and cultural heritage. A further question that arises, here, however, is whether the submerged part of the iceberg –the keel– while clearly caused by social silence and inhibition, is not also attributable to ignorance, which makes the problem of tackling the underreporting of GBV even more complex. Whatever the reasons for the underreporting of GBV, here our principal concern is quantifying the magnitude of cases of gender violence reported in the primary care system and in attempting to reconstruct the complete GBV iceberg. Reducing GBV is at the top of the policy agenda of many governments and organizations and the analysis of these policies and associated campaigns highlights the complexity of the issue and the data biases that beset any discussion of the problem ([8]). Several authors ([9, 10, 11, 12]) have undertaken studies of the legislation passed and public policies adopted to address GBV. Specifically, [13] conducted a descriptive-comparative study of health policies, plans, and protocols that proposed health actions targeting violence against women in Catalonia and Costa Rica between 2005 and 2011, and [14] highlights, on the basis of a large study conducted in India that gender-based police interventions (training, outreach and dedicated spaces staffed by female officers) led to a reduction in the underreporting of GBV. The Health Department in Catalonia was reported as putting the following strategies into effect: training actions in care services related to instances of male violence; promoting women’s groups and associations focused on GBV; implementing and evaluating health protocols; training for the treatment of sexual assault victims; care and prevention of female genital mutilation; and, the publication of materials for fostering community work. Given that these and other campaigns and policies have been implemented in recent years, it is interesting to assess their impact on the actual reporting of cases of GBV.

The fight against GBV and the efforts to promote gender equality are, likewise, currently at the top of the policy agendas of the Spanish and Catalan governments. Since the mid-2000s, Spain has implemented integrated programs to fight GBV, including the running of awareness campaigns, the creation of a Ministry for Equality and the establishment of specialized GBV courts ([15]). According to Spanish Organic Law 1/2004 on Integrated Protection Measures against Gender Violence, health professionals should be alert to the possibility of identifying cases of GBV when visiting patients ([16]). Primary care units should strive to manage these cases appropriately and form part of a multi-disciplinary response in coordination with other institutions and sectors. In acting against GBV, healthcare professionals play a leading role precisely because of their proximity to patients. In Catalonia, the Catalan Women’s Institute (*Institut Català de les Dones*), an organization attached to the Department of Equality and Feminism, designs, promotes, coordinates and evaluates the gender equality policies developed by the Regional Government (*Generalitat de Catalunya*). The Institute conducts GBV awareness training plans and intervention programs and provides both victims and the wider population with a range of resources and public services.

In Spain, few studies to date have attempted to evaluate the impact of policies or political interventions on the occurrence of episodes of GBV and, to the best of our knowledge, no analyses have been undertaken of the impact of non-political interventions, such as formative campaigns, on the GBV notification process. Here, in seeking to reconstruct the GBV iceberg, the importance of considering specific events and interventions that have an impact on GBV becomes apparent. Specifically, we evaluate the impact of two key events on the number of reported cases of GBV:

1. The COVID-19 pandemic and

2. The implementation of awareness campaigns among primary healthcare professionals, initiated in late 2019.

In March 2020, the state of emergency declared by the World Health Organization following the outbreak of the COVID-19 epidemic resulted in Spain, along with many other countries, ordering the mandatory confinement of the whole population from March 14th to May 1st, 2020, a lockdown that would be maintained, albeit not so rigorously, until June 24th, 2020. Confinement to the home was a notable risk factor for GBV as economic, employment, stress and mental health problems rose alarmingly and victims and aggressors were subjected to a prolonged period of cohabitation. A cross-sectional analysis conducted by [17] shows that the number of calls to Spain’s GBV helpline (dial 016) presented a marked increase between March and June 2020 compared to trends for the previous two years.

According to one news report ([18]), the number of registered cases in primary healthcare centers in Mexico during the COVID-19 pandemic does not reflect the actual number of GBV episodes, given that 96.4% of acts of sexual violence between July and December 2020 were not registered. Thus, we should not lose track of the possibility that even though an increase in GBV episodes was reported during the lockdown, these figures may also be subject to underreporting. Another current challenge that might be worsening the situation regarding GBV is climate change, as studied in [19].

In what follows, we use advanced statistical modelling techniques to calculate what lies beneath the tip of the GBV iceberg, that is, we seek to estimate the expected number of weekly registered cases of physical and sexual violence directed against women. To do this, we, first, describe our data sources for a specific metropolitan area of the Barcelona (Catalonia, Spain) health region and, second, develop our mathematical model for three specific scenarios. We then present the corresponding parameter estimates for the three simulations in each subarea of our health area. We find that awareness-raising activities targeting primary healthcare workers can greatly reduce underreporting but that, without additional activities, it could take Catalonia a further twenty years to reveal the whole of the GBV iceberg.

## 2 Methods

### 2.1 Data

The public health system in Catalonia is divided in seven health regions (*Regió Sanitària*):

- Regió Sanitària Alt Pirineu i Aran
- Regió Sanitària Lleida
- Regió Sanitària Camp de Tarragona
- Regió Sanitària Terres de l’Ebre
- Regió Sanitària Catalunya Central
- Regió Sanitària Girona
- Regió Sanitària Barcelona

The *Regió Sanitària Barcelona* is further divided into three metropolitan or city areas:

- Àmbit Metropolità Sud
- Àmbit Metropolità Nord
- Barcelona Ciutat

Each of these areas, in turn, is divided into a number of subareas, which are not identified here geographically so as to preserve patient anonymity. Our study draws on three sources of data: The weekly number of GBV cases registered in the *Àmbit Metropolità Nord* of the Barcelona health region between January 2010 and December 2021 in which the victim was a woman aged 16 or older; the total number of women aged 16 or over assigned to each of its six subareas (labelled A, B, C, D, E, and F for the purposes of our study); and the survey on violence against women conducted by the Spanish Ministry of Equality in 2019. The latter provides the only official statistics in Spain on the prevalence of violence against women, and is conducted every four years. The main objective of the 2019 survey was to determine the percentage of women aged 16 or over residing in Spain that have been the victims of any type of GBV. The interviews were conducted with a representative sample of 9,568 women. Surveys of this type provide us with a more realistic idea of the scale of GBV in Spain whether the episodes have been reported or not. Given that the interviews are anonymous, any social stigma and lack of access to resources and support systems should not constitute an obstacle to the correct quantification of GBV cases, and, thus, they offer a better approximation of the whole GBV iceberg in Spain. According to the survey, 31.73% of the women surveyed in the Barcelona area have suffered physical or sexual violence at some point in their lives, while only 15.65% of the victims attended, or considered that she should attend, a primary healthcare center as a result of the GBV suffered. On the basis of these two percentages and the number of women assigned to each subarea of the *Àmbit Metropolità Nord* in Barcelona (see Table 1), we calculated the expected number of weekly registered cases of GBV per area (further details on how these values are calculated are provided in the next section.)

**Table 1:**
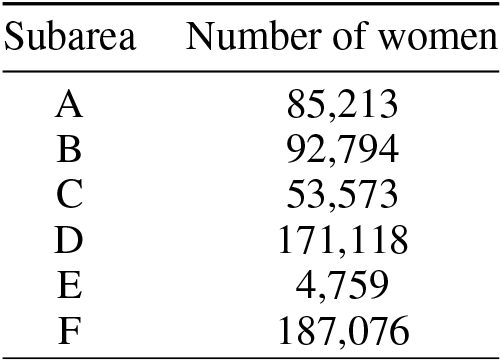
Number of women (aged 16 and over) assigned to each subarea.

| Subarea | Number of women |
| --- | --- |
| A | 85,213 |
| B | 92,794 |
| C | 53,573 |
| D | 171,118 |
| E | 4,759 |
| F | 187,076 |

It should be stressed that the *Àmbit Metropolità Nord* is one of the largest health areas in Catalonia, serving 1.5 million individuals of highly heterogeneous sociodemographic and socioeconomic characteristics, and that it includes densely populated municipalities, such as Santa Coloma de Gramanet (16,854.4 individuals per *km*^2^ in 2022, with one of the lowest per capita GDPs in Catalonia, 12.1 thousand euros in 2021, the latest value available) as well as rural municipalities, such as Tagamanent (7.8 individuals per *km*^2^ in 2022) and towns with a per capita GDP above the regional mean, the case of Granollers (38.9 thousand euros in 2021, while the Catalan mean was 31.6 thousand euros). As such, the results of this study are readily generalizable to the rest of Spain and even to other countries that operate similar public primary care systems.

### 2.2 Model

Let’s assume that the actual weekly number of GBV cases *X*_*t*_ follows a Poisson distribution with mean *λ*, which is increased by a factor of *β* in the mandatory confinement period (March 14th to June 24th, 2020), i.e., *E*(*X*_*t*_) = *λ*+*I*(*t*)·*β* where *I*(*t*) takes the value 1 if *t* falls within the mandatory confinement period and 0 otherwise.

The number of cases diagnosed within the public primary health care system, *Y*_*t*_, is just a part of the actual process *X*_*t*_, following a binomial distribution with parameters *q*_0_ and *X*_*t*_ from *t* = 1, …, *t*^*′*^ and with parameters *q*_*t*_ and *X*_*t*_ from *t* = *t*^*′*^ + 1, …, *n*, where *t*^*′*^ is the unknown change point at which awareness training for primary health care professionals starts impacting the reported weekly number of diagnoses and 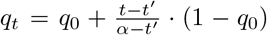 for *t > t*^*′*^.Additional details on the model structure can be found in the Supplementary Material.

It should be borne in mind that the number of GBV cases *X*_*t*_ is not directly available, as only the number of diagnosed cases *Y*_*t*_ is observed. Model (3) assumes that *Y*_*t*_ only reports a fraction *q*_*t*_ of the total number of GBV cases, and *q*_0_ can be interpreted as the proportion of reported cases in the first part of the series while *q*_*t*_ grows towards 1 as a consequence of the awareness training (as it contributes to reducing the underreporting). Additionally, an alternative exponential modelling of *q*_*t*_ as 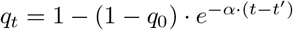 was considered and compared to the previously described approach in terms of the deviance information criterion (DIC) (see Table 3). The DIC is an estimator of prediction error and relative quality of statistical models for a given set of data, and it is commonly used in Bayesian model selection problems where the posterior distributions of the models have been obtained by Markov chain Monte Carlo (MCMC) simulation as a means to compare models. The DIC is defined as:

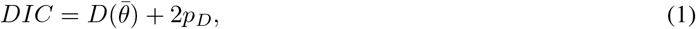

where *D*(*θ*) = −2 · *log*(*p*(*y* | *θ*)) + *C*, and *y* are the observations, 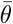 the expectation of the parameters vector *θ* and *C* a constant that cancels out in all calculations that compare different models, and which does not, therefore, need to be known. In line with [20], *p*_*D*_ is computed as 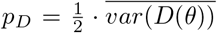. When comparing different models’ DIC, the preferred model is the one with the lowest value. Here, each subarea was modelled according to the best fitting approach.

It should also be noted that for the linear modelling of *q*_*t*_, *α* is the moment when *q*_*α*_ = 1, i.e., the registered and observed processes coincide. In the case of the exponential modelling of *q*_*t*_, *α* is proportional to the speed at which the underreporting issue disappears as a consequence of the training activity, so it is also related to the moment in time when all cases are registered. All the parameters (*q*_0_, *λ, β, α* and *t*^*′*^) are estimated by Gibbs sampling, aimed at constructing a Markov chain with values converging to a target distribution, using the *R2jags* package [21], taking appropriate priors based on the information available. In order to avoid non-identifiability of the model (3), a hierarchical Bayesian modelling approach was used, with each subarea as a level, assuming that the actual average number of GBV cases in each subarea during the non-confinement period (parameter *λ*) follows a normal prior distribution with standard deviation *σ* = 1 and with a mean based on three realistic scenarios described below, leading to the expected weekly average number of cases detailed in Table 2:

**Table 2:** Expected weekly average number of diagnosed GBV cases in each subarea by scenario.

| Subarea | Scenario 1 | Scenario 2 | Scenario 3 |
| --- | --- | --- | --- |
| A | 7 | 21 | 42 |
| B | 7 | 23 | 46 |
| C | 4 | 13 | 26 |
| D | 13 | 42 | 84 |
| E | 1<sup>a</sup> | 1 | 2 |
| F | 14 | 46 | 92 |

<sup>a</sup>The expected average number of diagnosed GBV cases based on the Spanish GBV survey for subarea E is zero, owing in all probability to the low number of women assigned to this subarea. However, we opt to consider the expected weekly mean number of diagnosed cases as 1 so as to be able to produce the model estimations for this subarea.

1. Scenario 1: The expected situation according to the results of the macro survey conducted by the Spanish Ministry of Equality, i.e., around 32% of women in the Barcelona area have suffered physical or sexual GBV at some point in their lives, and only around 16% of the victims sought medical care after suffering such an episode (excluding hospital admissions). It should be stressed that this is the most conservative scenario, as it assumes that the results of the survey are not underestimating the prevalence of GBV cases and subsequent usage of health services, which is dubious to say the least. In this scenario, the weekly incidence per 100,000 women is around 8 cases.

2. Scenario 2: Assuming again that around 32% of women in the Barcelona area have suffered physical or sexual GBV at some point in their lives, but considering that 50% of the victims seek medical care after suffering such an episode (excluding hospital admissions). In this scenario, the weekly incidence per 100,000 women is around 25 cases.

3. Scenario 3: Assuming again that around 32% of women in the Barcelona area have suffered physical or sexual GBV at some point in their lives, but considering that 90% of the victims seek medical care after suffering such an episode (excluding hospital admissions). This scenario is especially interesting from a preventive perspective, as it is known that 90% of the population attends a primary health center at least once every two years and, therefore, this is scenario considers primary care as the reference system for cases of GBV. In this scenario, the weekly incidence per 100,000 women is around 45 cases.

The values in Table 2 have been computed taking into account the assumptions of each scenario and the number of assigned women in each subarea. For instance, in the case of scenario 1 in subarea A, the figure of 7 expected cases per week is obtained by considering the nearest integer to 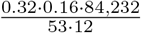, given that we have data for 53 weeks in each of the 12 years making up the period of study.

Once the parameters have been estimated, the most likely process can be reconstructed taking into account that *Y*_*i*_|*X*_*i*_ ∼ *Binom*(*x*_*i*_, *q*_*t*_). At each time *t* with *j* reported cases, the most likely number of GBV cases is the value *ν* that maximizes the probability

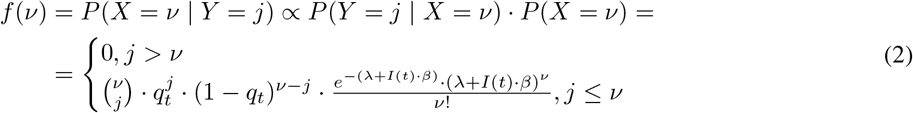

Additionally, we considered an exhaustive simulation study reproducing the described structure with different parameter values has been conducted in order to assess whether the original values can be recovered by using this estimation method and to evaluate the model performance, both for the linear and exponential modelling of *q*_*t*_. The results of the simulation study are presented in the Supplementary Material, summarized in Table S1.

## 3 Results

### 3.1 Catalonia Primary Healthcare System

#### 3.1.1 Scenario 1: Baseline average number of weekly cases based on the macro GBV survey (2019)

All the results reported here correspond to exponential *q*_*t*_, our preferred assumption in all six subareas, as shown in Table 3.

**Table 3:** Deviance information criterion (DIC) corresponding to the linear and exponential modelling of *q*_*t*_ in each subarea.

| Subarea | Linear | Exponential |
| --- | --- | --- |
| A | 1987.3 | 1979.7 |
| B | 2293.6 | 2289.5 |
| C | 1361.9 | 1359.0 |
| D | 3266.5 | 3251.5 |
| E | 2051.1 | 1830.2 |
| F | 2596.7 | 2572.4 |

The first question we need to consider is whether the theoretical model described is able to reproduce the process observed accurately. A simulation based on the theoretical model that uses the estimated parameters corresponding to scenario 1 for each subarea is shown in Figure 1 together with the processes observed. It is evident that the simulated processes are able to capture the variability of the observations accurately in all six subareas.

**Figure 1:**
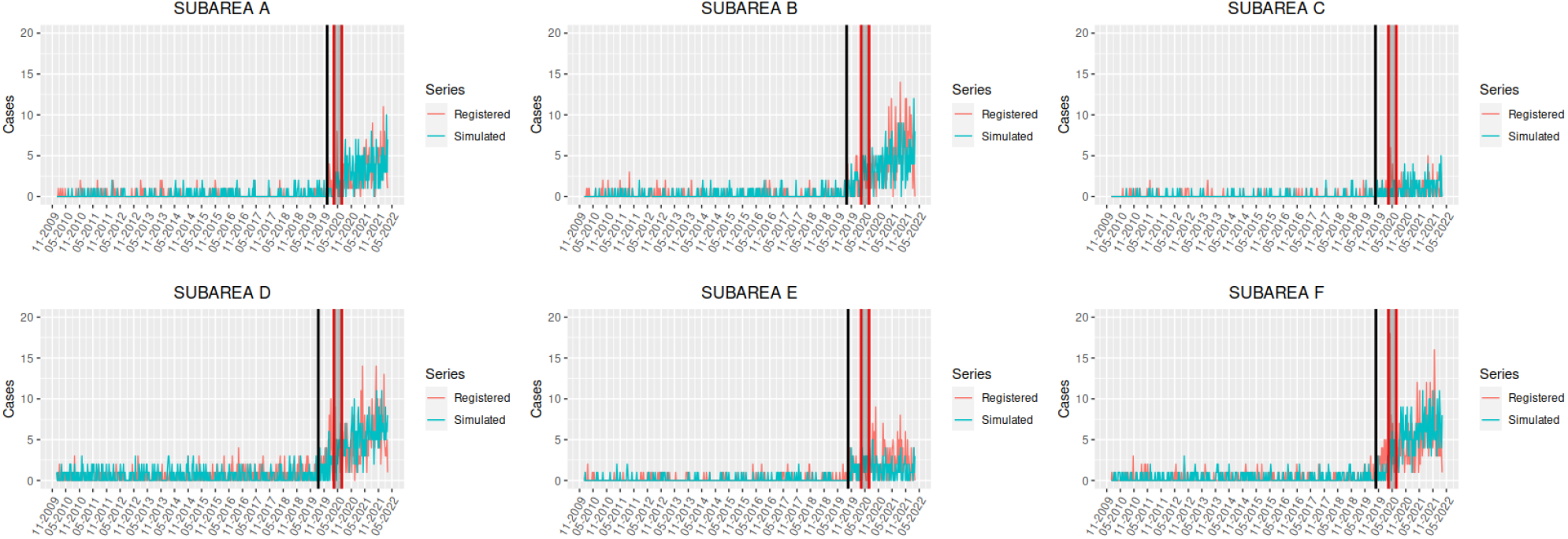
Evolution of the weekly number of GBV diagnoses in each subarea of the *Àmbit Metropolità Nord* (Catalonia, Spain) (in red) and estimated observed process according to the theoretical model (in green) for scenario 1. The solid black vertical line indicates the moment when training starts to impact the underreporting process, while the area between the red vertical lines represents the confinement period.

The analogous figures for scenarios 2 and 3 are reported in the Supplementary Material (Figures 11 and 12, respectively). Table 4 presents a summary of the parameter estimates in each subarea. It is evident that the underreporting at the beginning of the period is severe (median *q*_0_ ranging from 0.018 to 0.103), and that the impact of the awareness training for professionals contributes to reducing underreporting from very early dates, although the actual number of cases will not be fully registered until 2028-11-03, 2026-02-20, 2041-05-31, 2033-11-04, 2019-11-22, 2033-01-28 for subareas A, B, C, D, E and F, respectively, considering that these are the dates when *q*_*t*_ rises above 0.99 in each subarea. It should be borne in mind that only 4,695 women aged 16 or older are assigned to subarea E, while the other areas provide healthcare services to 84,232, 91,639, 52,968, 168,783 and 184,594 women respectively, and it therefore seems reasonable that the real number of cases is already being registered in this subarea.

**Table 4:**
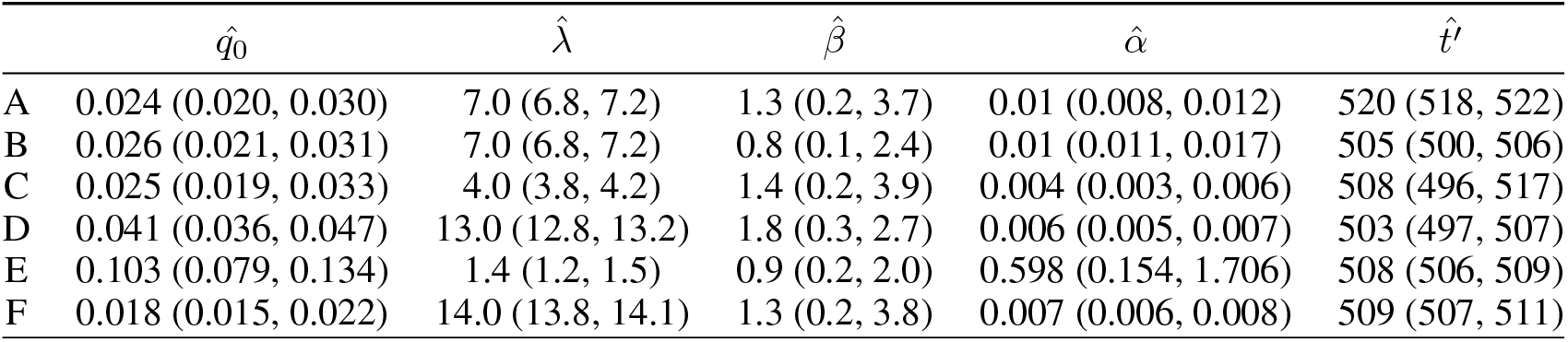
Parameter estimates (posterior median and 95% credible interval) for each subarea under scenario 1.

| | $\hat{q}_0$ | $\hat{\lambda}$ | $\hat{\beta}$ | $\hat{\alpha}$ | $\hat{t}'$ |
| --- | --- | --- | --- | --- | --- |
| A | 0.024 (0.020, 0.030) | 7.0 (6.8, 7.2) | 1.3 (0.2, 3.7) | 0.01 (0.008, 0.012) | 520 (518, 522) |
| B | 0.026 (0.021, 0.031) | 7.0 (6.8, 7.2) | 0.8 (0.1, 2.4) | 0.01 (0.011, 0.017) | 505 (500, 506) |
| C | 0.025 (0.019, 0.033) | 4.0 (3.8, 4.2) | 1.4 (0.2, 3.9) | 0.004 (0.003, 0.006) | 508 (496, 517) |
| D | 0.041 (0.036, 0.047) | 13.0 (12.8, 13.2) | 1.8 (0.3, 2.7) | 0.006 (0.005, 0.007) | 503 (497, 507) |
| E | 0.103 (0.079, 0.134) | 1.4 (1.2, 1.5) | 0.9 (0.2, 2.0) | 0.598 (0.154, 1.706) | 508 (506, 509) |
| F | 0.018 (0.015, 0.022) | 14.0 (13.8, 14.1) | 1.3 (0.2, 3.8) | 0.007 (0.006, 0.008) | 509 (507, 511) |

The evolution of the phenomenon in each subarea is shown in Figure 2, together with a reconstruction of the most likely actual process according to Equation (2) and taking the median of the estimated posterior distribution for each parameter.

**Figure 2:**
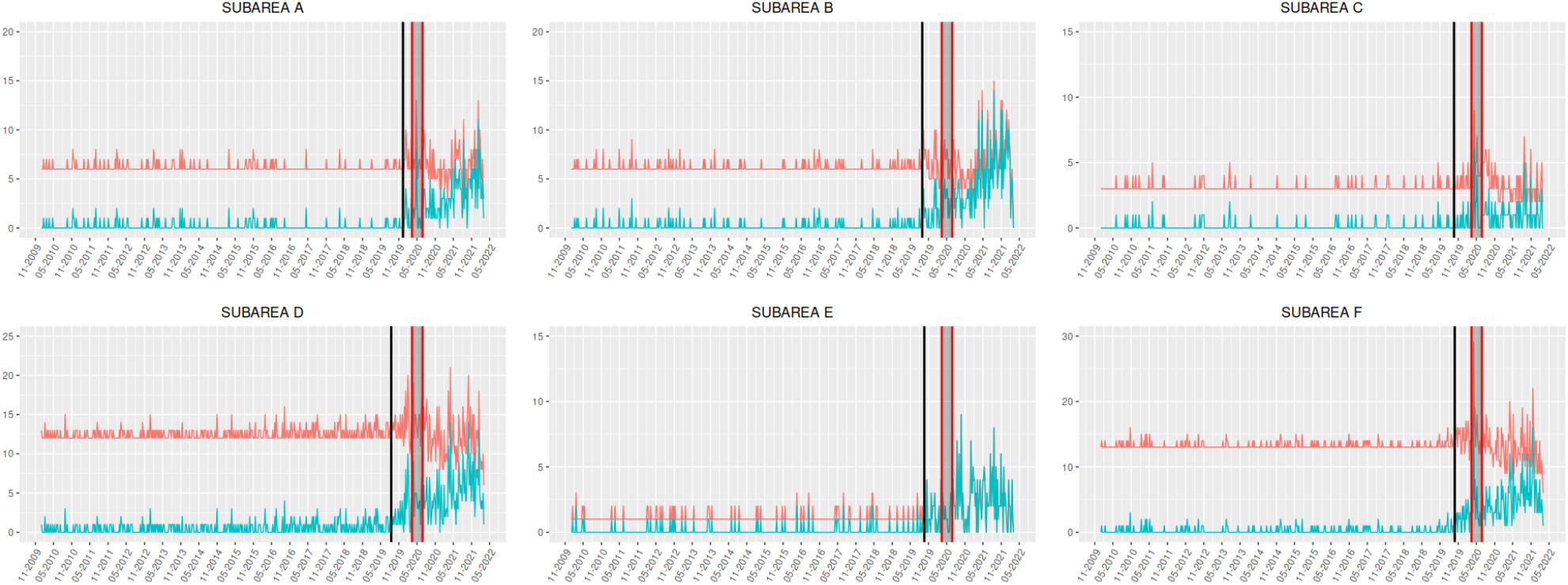
Evolution of the weekly number of GBV diagnoses in each subarea of the *Àmbit Metropolità Nord* (Catalonia, Spain) (in green) and a reconstruction of the most likely process according to Eq. (2) (in red) for scenario 1. The solid black vertical line indicates the moment when training starts to impact the underreporting process, while the area between the red vertical lines represents the confinement period.

Figure 3 shows the impact of the training intervention on each subarea through the evolution of 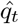.

**Figure 3:**
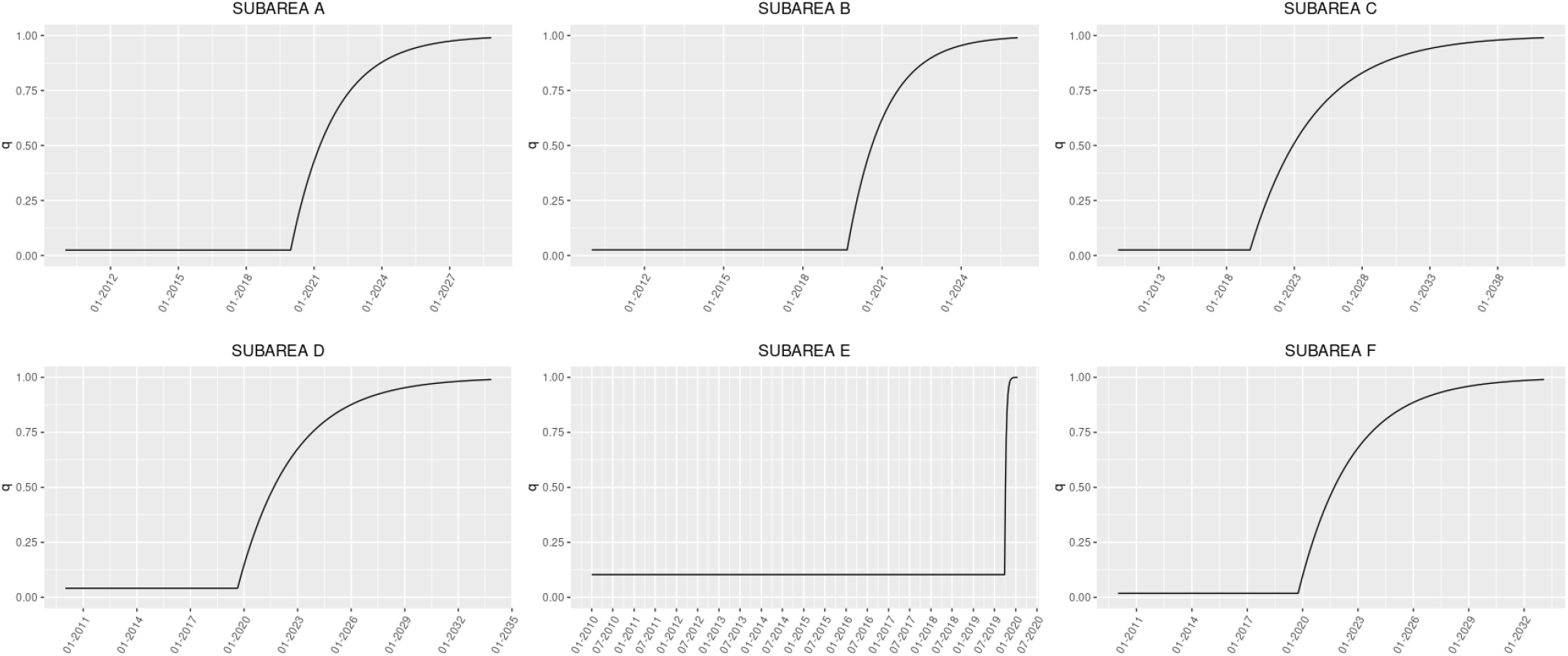
Evolution of 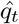 in each subarea according to scenario 1.

By adding the observed and estimated GBV events within the time period considered for each subarea, the percentage of cases registered (over the estimated total) can also be computed, as shown in Figure 4.

**Figure 4:**
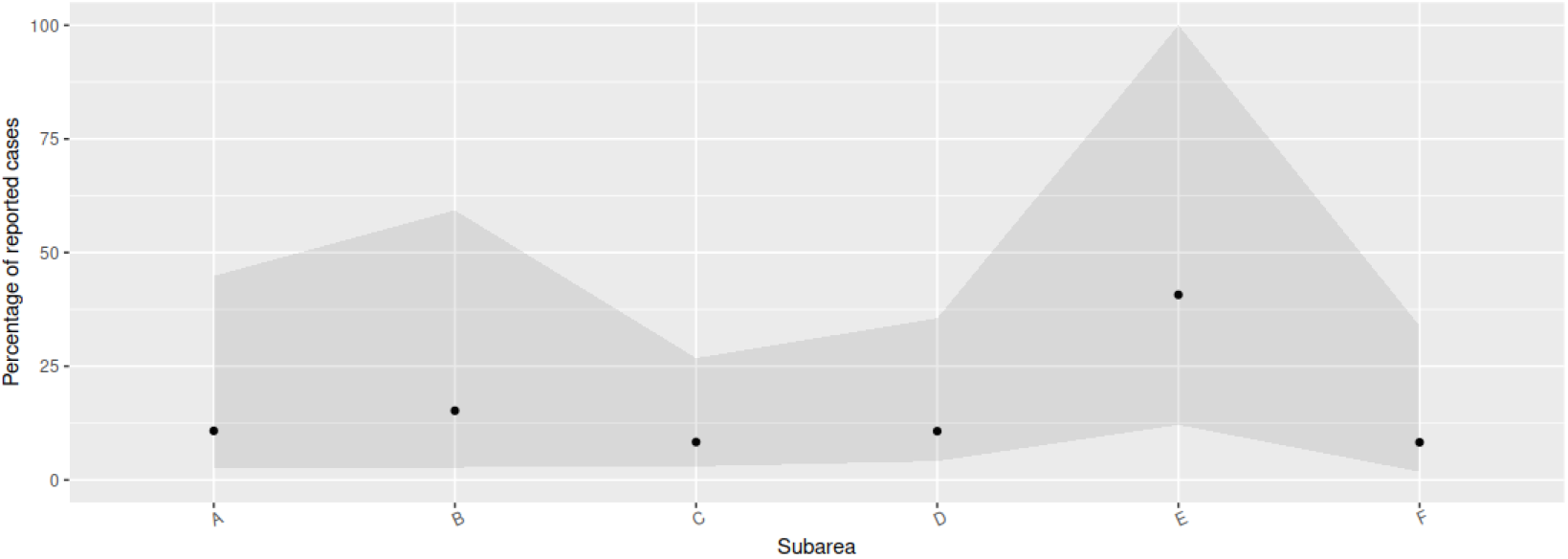
Percentage of cases registered over the estimated actual number of cases in each subarea cumulated over the whole time period (black point). The grey shaded area represents the percentage of cases registered before (minimum) and after (maximum) the training activity according to scenario 1.

In each case, we ran five MCMC chains and 50,000 iterations, and convergence was reached for all parameters without any particular patterns presenting themselves in the trace plots, as can be seen in Figures 13 to 18 in the Supplementary Material.

#### 3.1.2 Scenario 2: 50% of women seek medical care (excluding hospital admissions) after suffering an episode of GBV

As in scenario 1, the DIC for the exponential *q*_*t*_ were slightly lower, so the results reported here correspond to this, our preferred, assumption. Assuming, as in the previous scenario, that around 32% of women have suffered a GBV episode at least once in their lifetime, but here that 50% of the victims seek medical care after suffering such an episode (excluding hospital admissions), we present the corresponding parameter estimates for each subarea are shown in Table 5. As in scenario 1, underreporting at the beginning of the period is severe (median *q*_0_ ranging from 0.006 to 0.103), and the impact of the awareness training for professionals contributes to reducing the underreporting from early dates, although the actual number of cases will not be fully registered until 2055-03-19, 2051-05-12, 2106-06-11, 2080-08-09, 2019-11-22, 2076-09-25 for subareas A, B, C, D, E and F, respectively.

**Table 5:** Parameter estimates (posterior median and 95% credible interval) for each subarea under scenario 2.

| | $\hat{q}_0$ | $\hat{\lambda}$ | $\hat{\beta}$ | $\hat{\alpha}$ | $\hat{t}'$ |
| --- | --- | --- | --- | --- | --- |
| A | 0.008 (0.007, 0.01) | 21.0 (20.8, 21.2) | 2.0 (0.3, 6.0) | 0.002 (0.002, 0.003) | 520 (518, 522) |
| B | 0.008 (0.006, 0.01) | 23.0 (22.8, 23.2) | 1.5 (0.2, 4.7) | 0.003 (0.002, 0.003) | 504 (500, 506) |
| C | 0.008 (0.006, 0.01) | 13.0 (12.8, 13.2) | 1.9 (0.3, 5.7) | 0.001 (0.001, 0.001) | 507 (484, 516) |
| D | 0.013 (0.011, 0.015) | 42.0 (41.8, 42.1) | 2.1 (0.3, 6.6) | 0.001 (0.001, 0.002) | 501 (495, 506) |
| E | 0.103 (0.079, 0.134) | 1.4 (1.2, 1.5) | 0.9 (0.2, 1.3) | 0.598 (0.154, 1.706) | 508 (506, 509) |
| F | 0.006 (0.005, 0.007) | 46.0 (45.8, 46.2) | 1.8 (0.3, 5.7) | 0.002 (0.001, 0.002) | 509 (503, 510) |

The evolution of the phenomenon in each subarea is shown in Figure 5, together with a reconstruction of the most likely actual process according to Equation (2).

**Figure 5:**
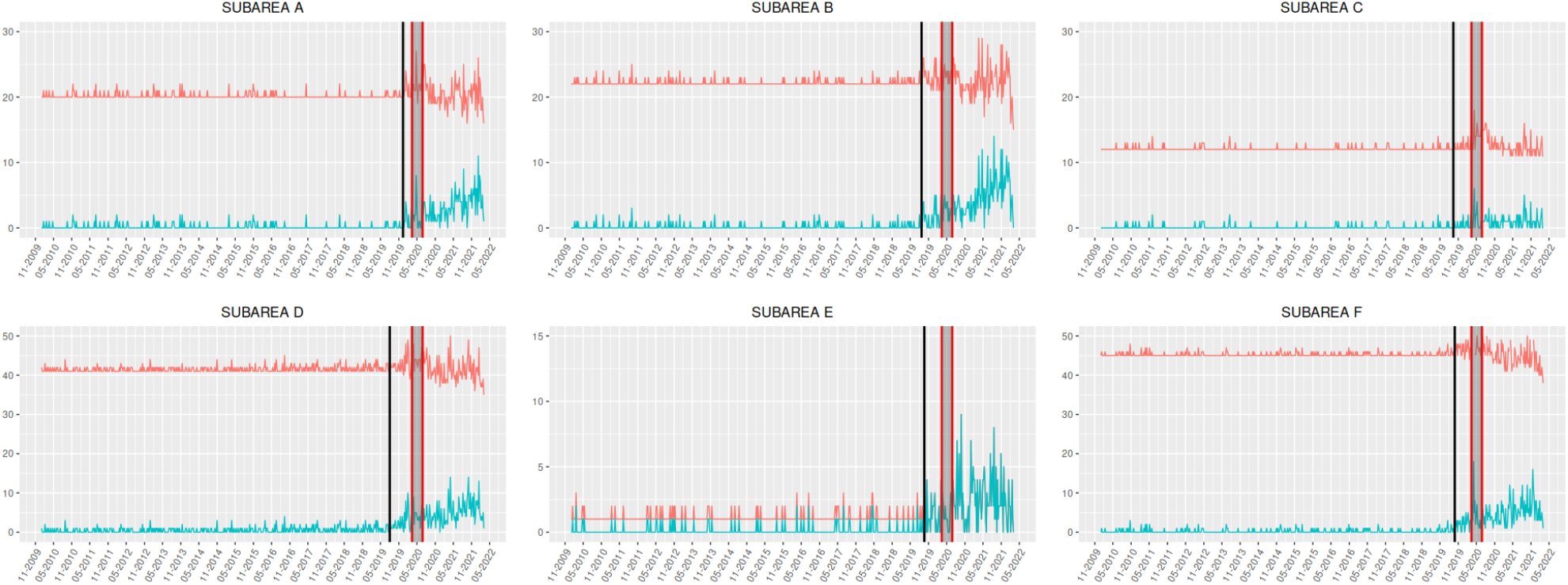
Evolution of the weekly number of GBV diagnoses in each subarea of the *Àmbit Metropolità Nord* (Catalonia, Spain) (in green) and a reconstruction of the most likely process according to Eq. (2) (in red) for scenario 2. The solid black vertical line indicates the moment when training starts to impact the underreporting process, while the area between the red vertical lines represents the confinement period.

Figure 6 shows the impact of the training intervention on each subarea through the evolution of 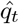 under scenario 2.

**Figure 6:**
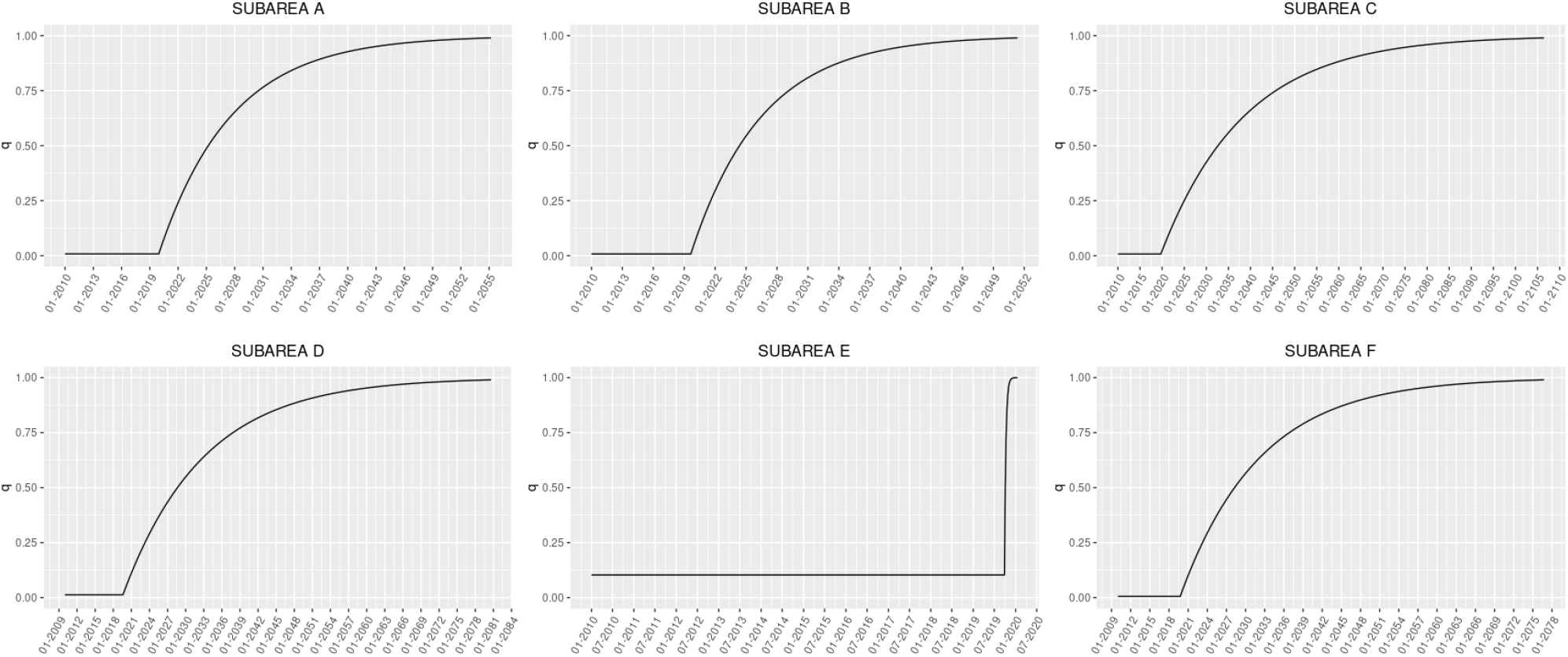
Evolution of 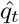 in each subarea according to scenario 2.

By adding the observed and estimated GBV events within the time period considered in each subarea, the percentage of cases registered (over the estimated total) can also be computed, as shown in Figure 7.

**Figure 7:**
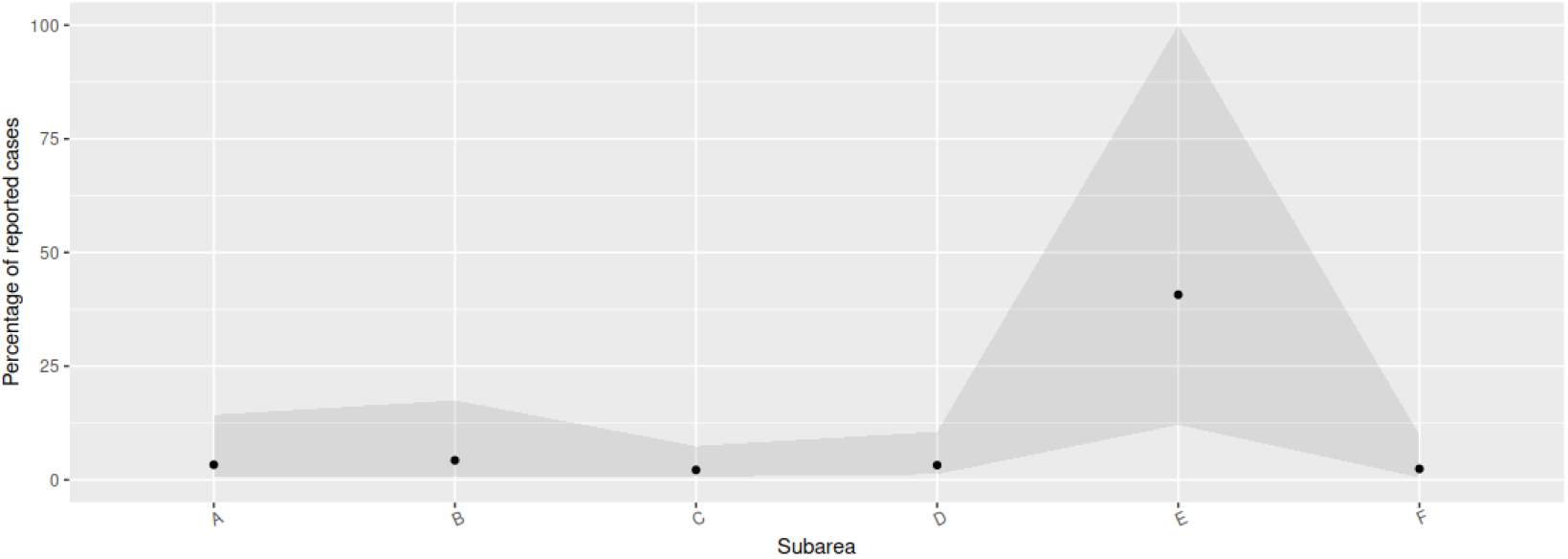
Percentage of cases registered over the estimated actual number of cases in each subarea cumulated over the whole time period (black point). The grey shaded area represents the percentage of cases registered before (minimum) and after (maximum) the training activity according to scenario 2.

#### 3.1.3 Scenario 3: 90% of women seek medical care (excluding hospital admissions) after suffering an episode of GBV

As in scenarios 1 and 2, the DIC for exponential *q*_*t*_ were slightly lower in this case, so the results reported here correspond to this, our preferred, assumption. Assuming, as in the other scenarios, that around 32% of women have suffered a GBV episode at least once in their lifetime, but here that 90% of the victims seek for medical care after suffering such an episode (excluding hospital admissions), we present the corresponding parameter estimates for each subarea in Table 6. As in scenarios 1 and 2, the underreporting at the beginning of the period is severe (median *q*_0_ ranging from 0.003 to 0.103), and the impact of the awareness training for professionals contributes to reducing the underreporting from very early dates, although the actual number of cases will not be fully registered until 2094-11-05, 2087-09-19, 2200-06-20, 2148-11-29, 2019-11-22, 2140-08-19 for subareas A, B, C, D, E and F, respectively.

**Table 6:** Parameter estimates (posterior median and 95% credible interval) for each subarea under scenario 3.

| | $\hat{q}_0$ | $\hat{\lambda}$ | $\hat{\beta}$ | $\hat{\alpha}$ | $\hat{t}'$ |
| --- | --- | --- | --- | --- | --- |
| A | 0.004 (0.003, 0.005) | 42.0 (41.8, 42.2) | 1.9 (0.3, 6.2) | 0.001 (0.001, 0.001) | 520 (518, 522) |
| B | 0.004 (0.003, 0.005) | 46.0 (45.8, 46.2) | 1.6 (0.2, 5.3) | 0.001 (0.001, 0.001) | 504 (500, 506) |
| C | 0.004 (0.003, 0.005) | 26.0 (25.8, 26.2) | 1.8 (0.3, 5.9) | 0.000 (0.000, 0.001) | 507 (483, 515) |
| D | 0.006 (0.006, 0.007) | 84.0 (83.8, 84.2) | 1.9 (0.3, 6.3) | 0.001 (0.001, 0.001) | 501 (495, 505) |
| E | 0.103 (0.079, 0.134) | 1.4 (1.2, 1.5) | 0.9 (0.2, 2.0) | 0.598 (0.154, 1.706) | 508 (506, 509) |
| F | 0.003 (0.002, 0.003) | 92.0 (91.8, 92.2) | 1.8 (0.3, 5.8) | 0.001 (0.001, 0.001) | 509 (503, 510) |

The evolution of the phenomenon in each subarea is shown in Figure 8, together with a reconstruction of the most likely actual process according to Equation (2) under this scenario.

**Figure 8:**
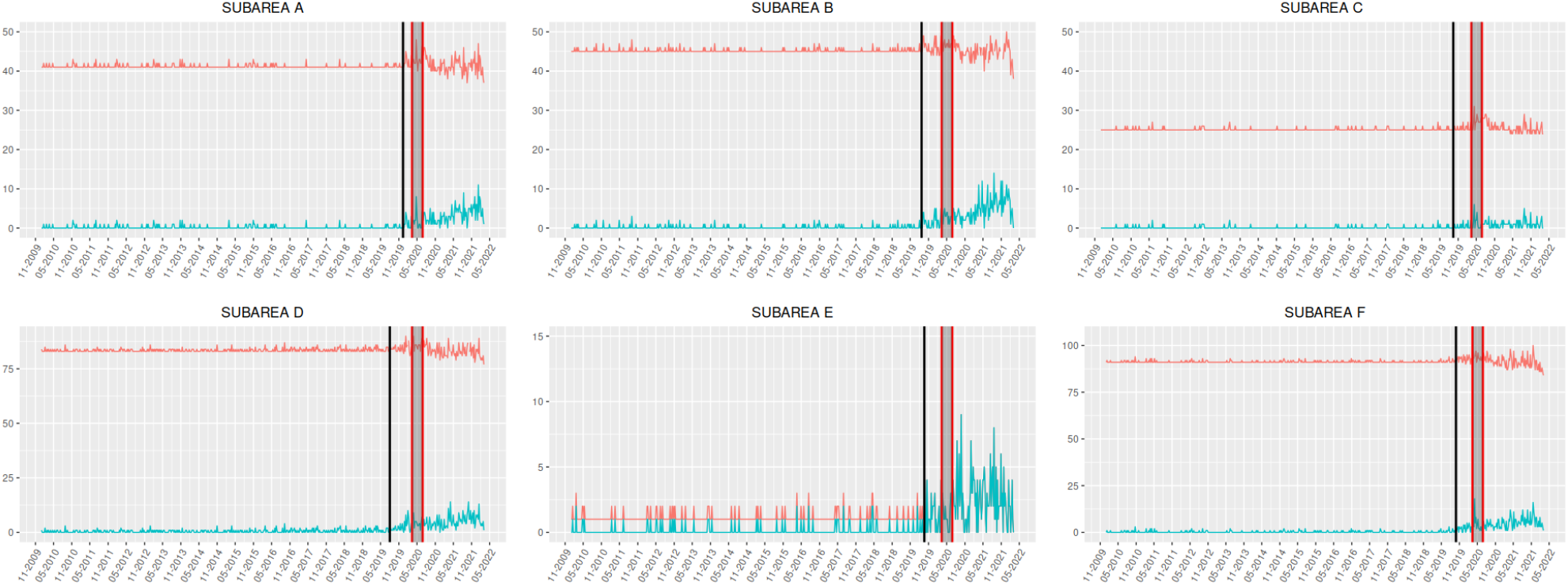
Evolution of the weekly number of GBV diagnoses in each subarea of the *Àmbit Metropolità Nord* (Catalonia, Spain) (in green) and a reconstruction of the most likely process according to Eq. (2) (in red) for scenario 3. The solid black vertical line indicates the moment when training starts to impact the underreporting process, while the area between the red vertical lines represents the confinement period.

Figure 9 shows the impact of the training intervention on each subarea through the evolution of 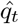 under scenario 3.

**Figure 9:**
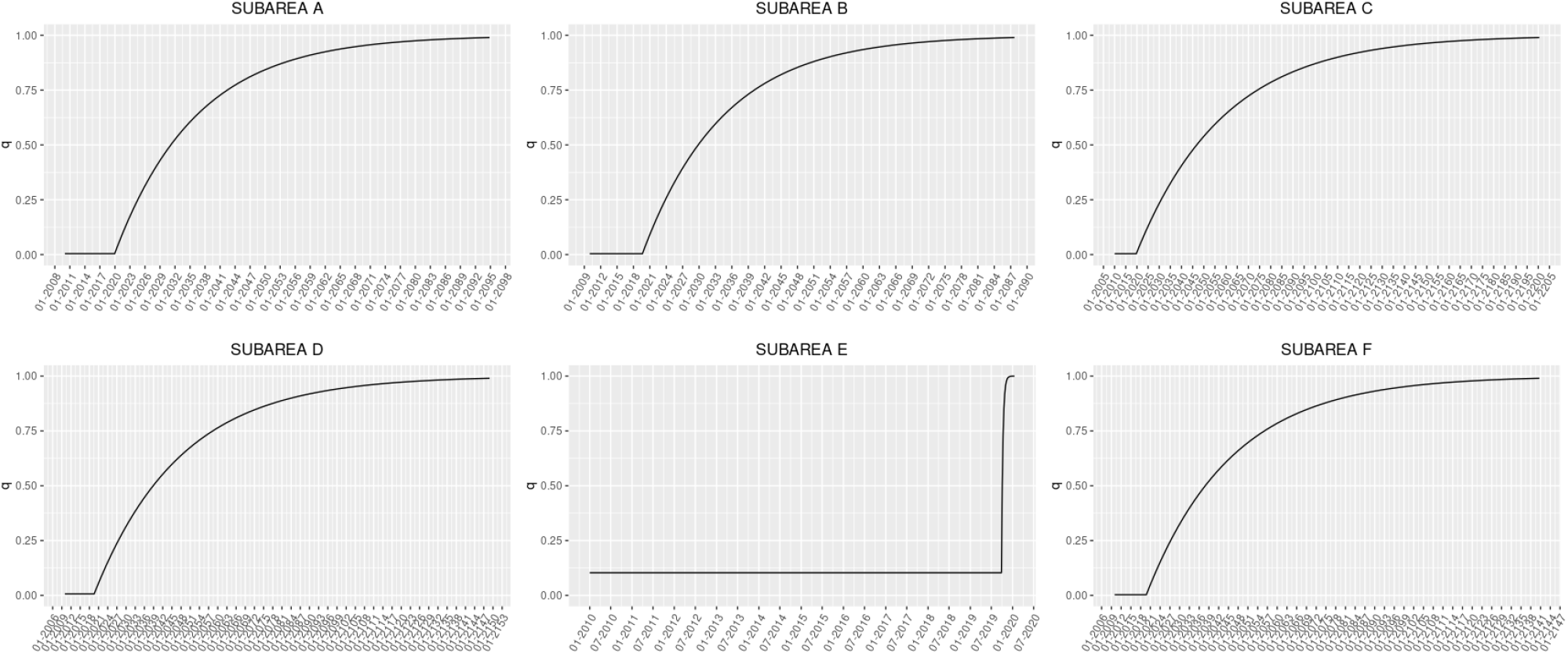
Evolution of 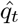 in each subarea according to scenario 3.

By adding the observed and estimated GBV events within the time period considered in each subarea, the percentage of cases registered (over the estimated total) can also be computed, as shown in Figure 10.

**Figure 10:**
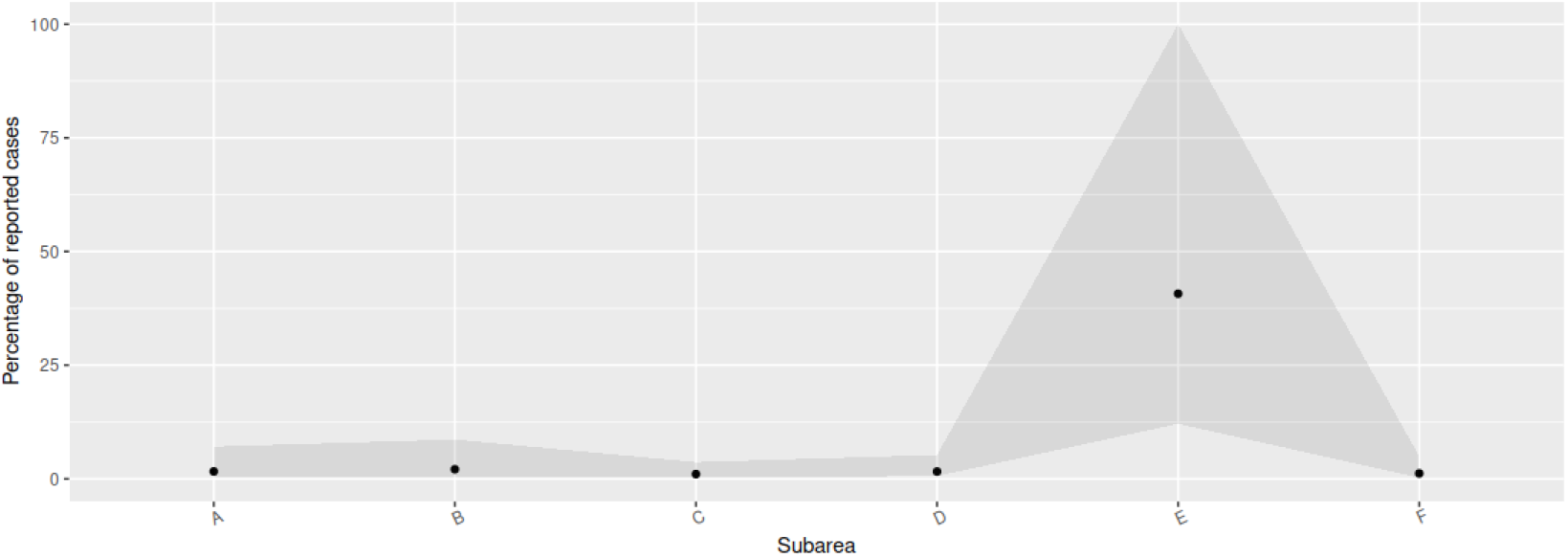
Percentage of cases registered over the estimated actual number of cases in each subarea cumulated over the whole time period (black point). The grey shaded area represents the percentage of cases registered before (minimum) and after (maximum) the training activity according to the scenario 3.

**Figure 11:**
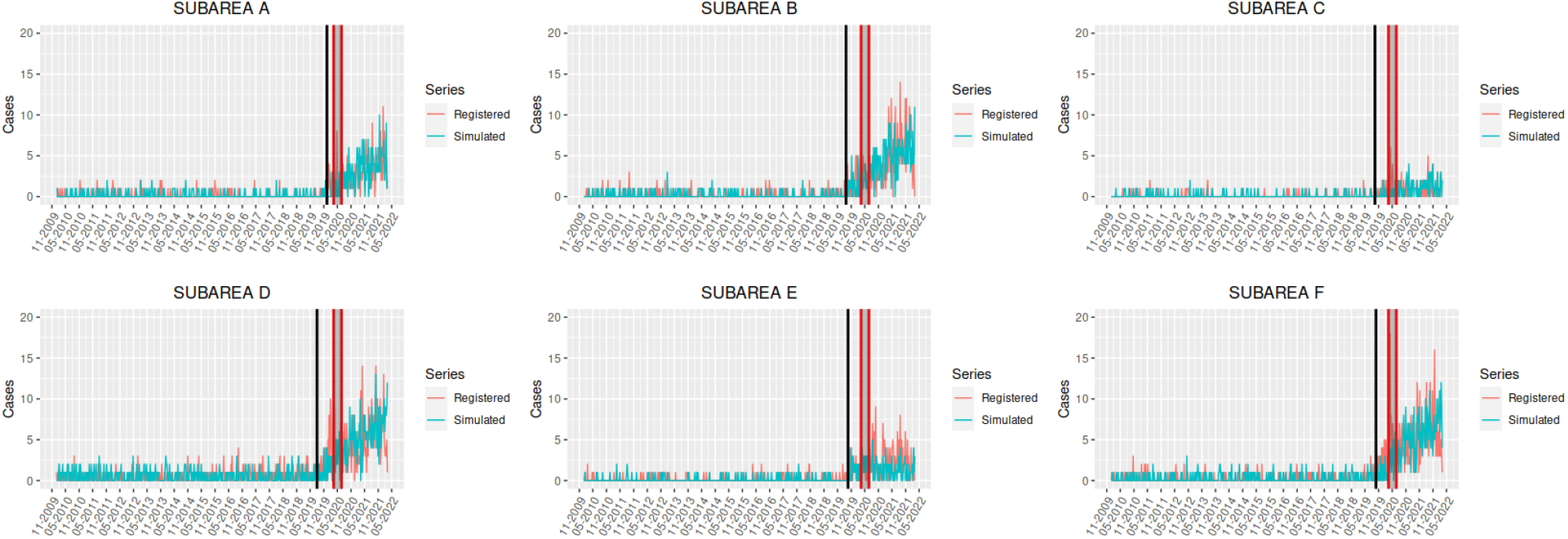
Evolution of the weekly number of GBV diagnoses in each subarea of the *Àmbit Metropolità Nord* (Catalonia, Spain) (in red) and estimated observed process according to the theoretical model (in green) for scenario 2. The solid black vertical line indicates the moment when training starts to impact the underreporting process, while the area between the red vertical lines represents the confinement period.

**Figure 12:**
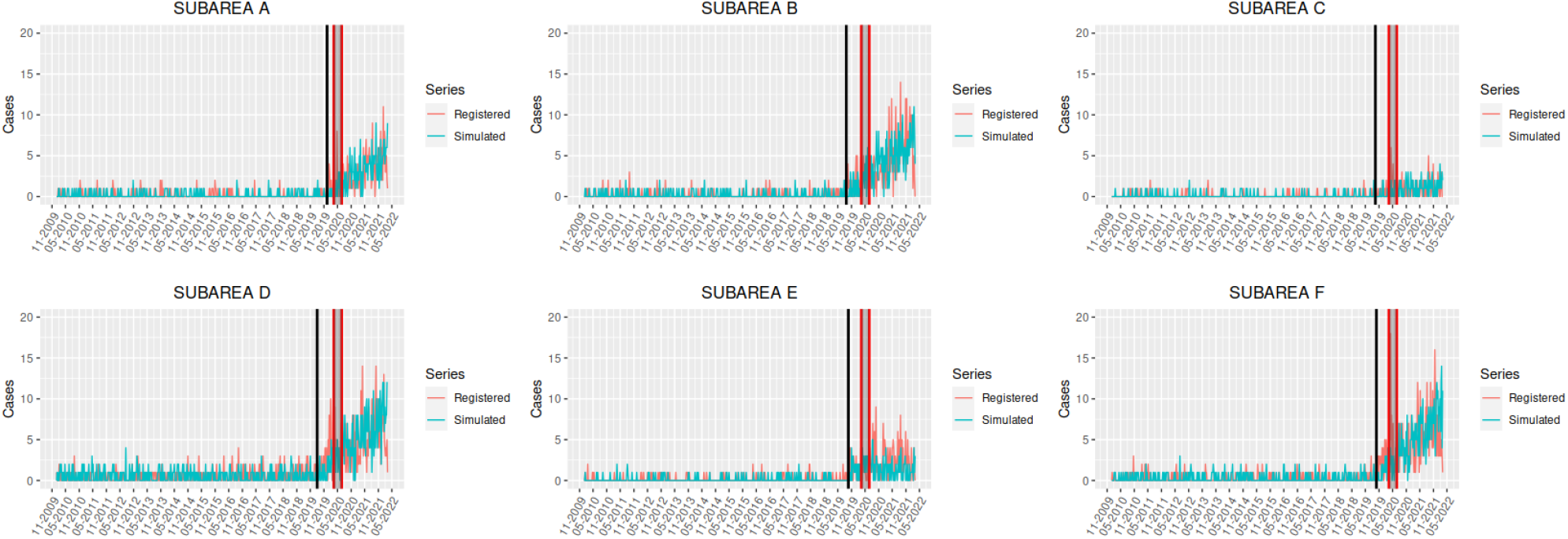
Evolution of the weekly number of GBV diagnoses in each subarea of the *Àmbit Metropolità Nord* (Catalonia, Spain) (in red) and estimated observed process according to the theoretical model (in green) for scenario 3. The solid black vertical line indicates the moment when training starts to impact the underreporting process, while the area between the red vertical lines represents the confinement period.

**Figure 13:**
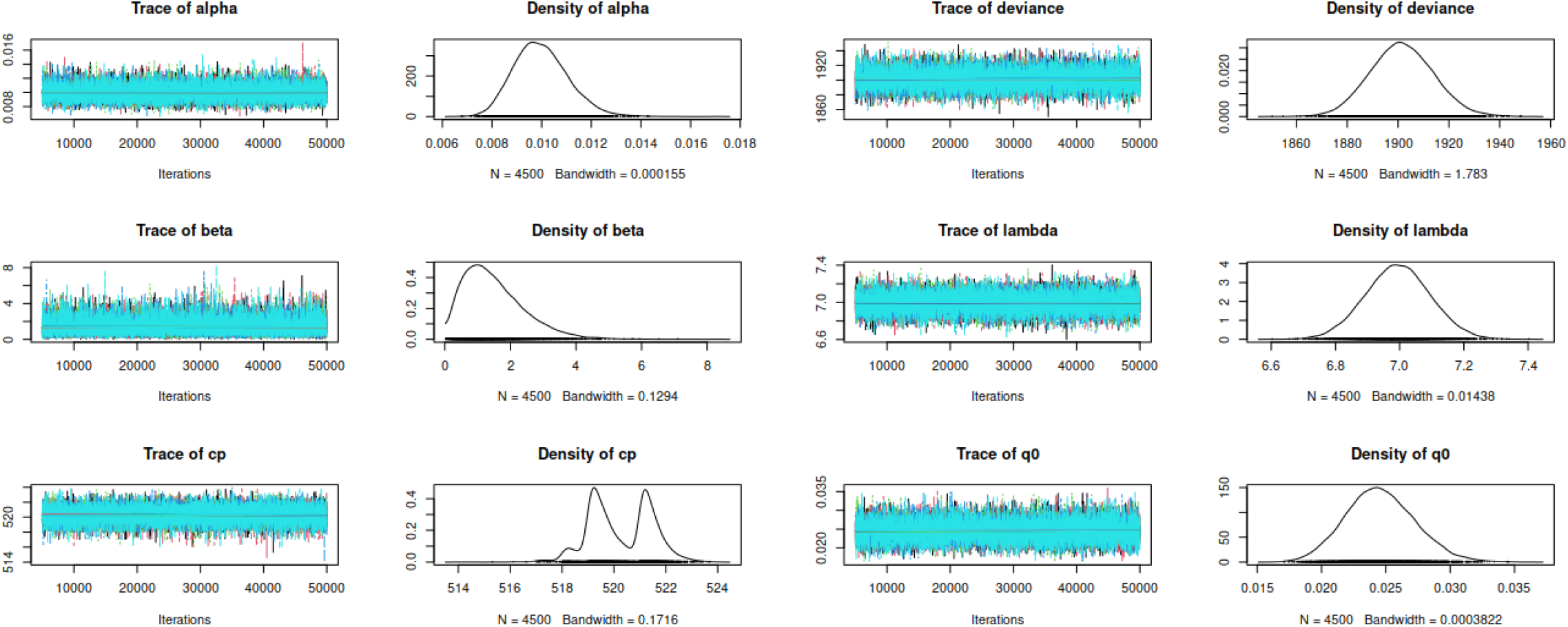
MCMC diagnostics: Chain convergence and posterior density functions for subarea A under scenario 1.

**Figure 14:**
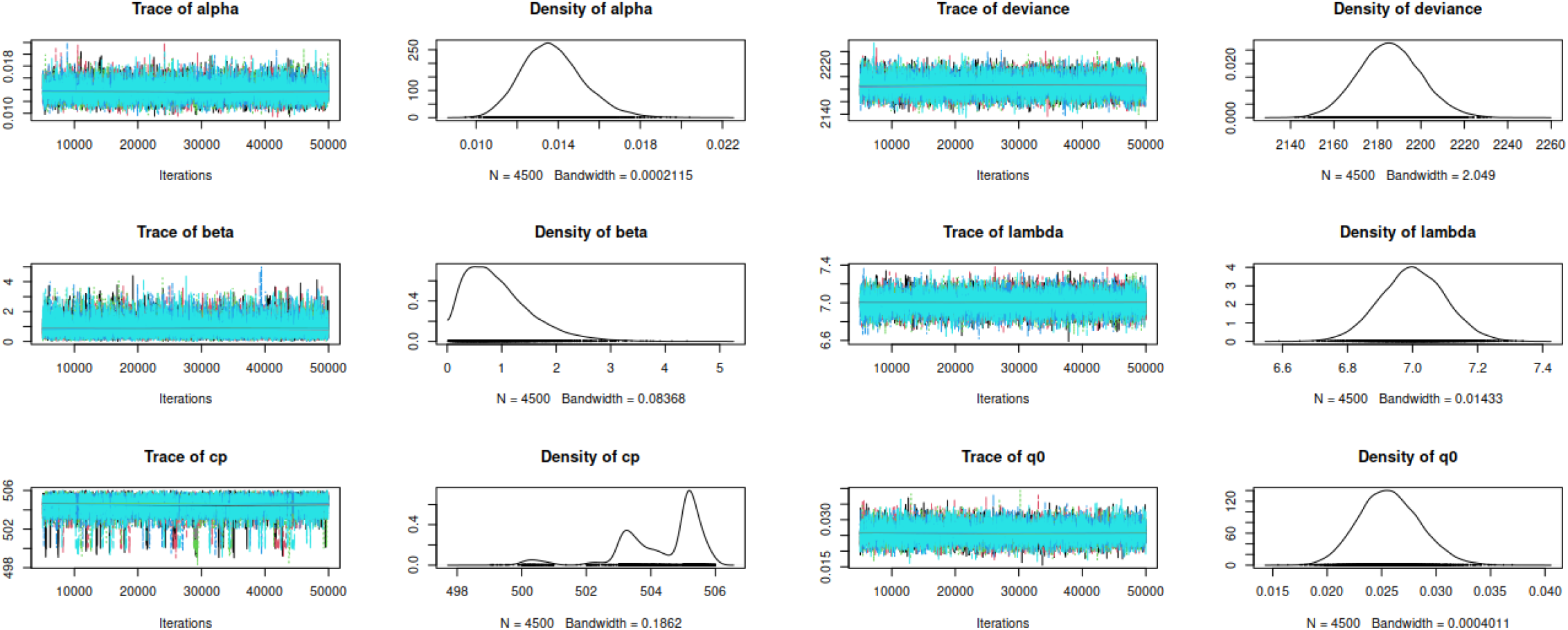
MCMC diagnostics: Chain convergence and posterior density functions for subarea B under scenario 1.

**Figure 15:**
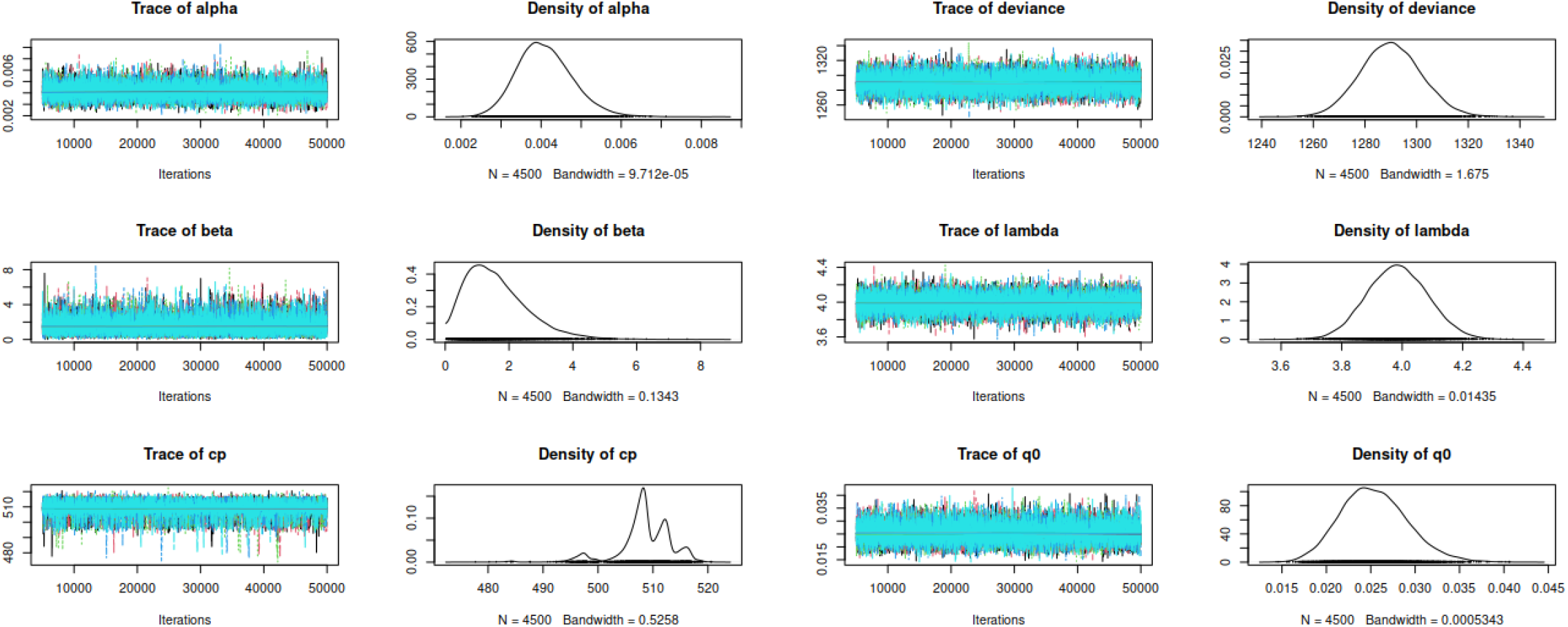
MCMC diagnostics: Chain convergence and posterior density functions for subarea C under scenario 1.

**Figure 16:**
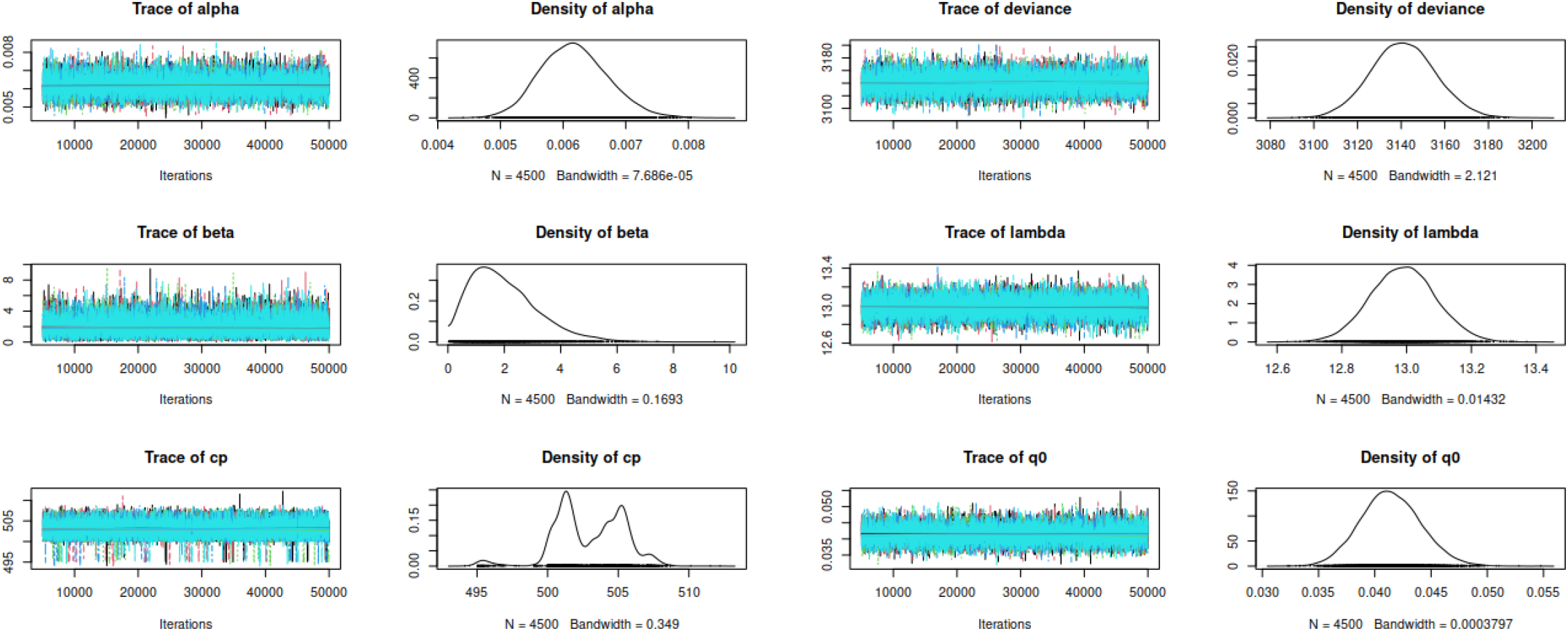
MCMC diagnostics: Chain convergence and posterior density functions for subarea D under scenario 1.

**Figure 17:**
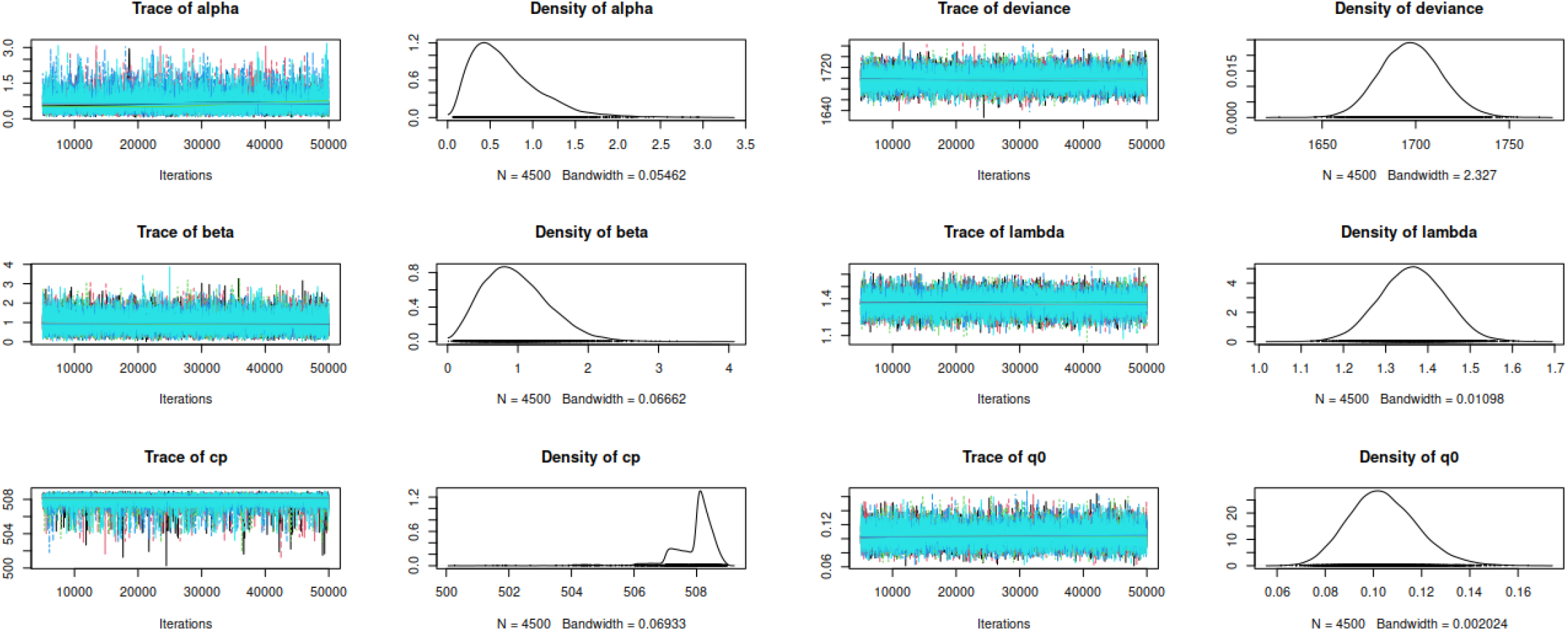
MCMC diagnostics: Chain convergence and posterior density functions for subarea E under scenario 1.

**Figure 18:**
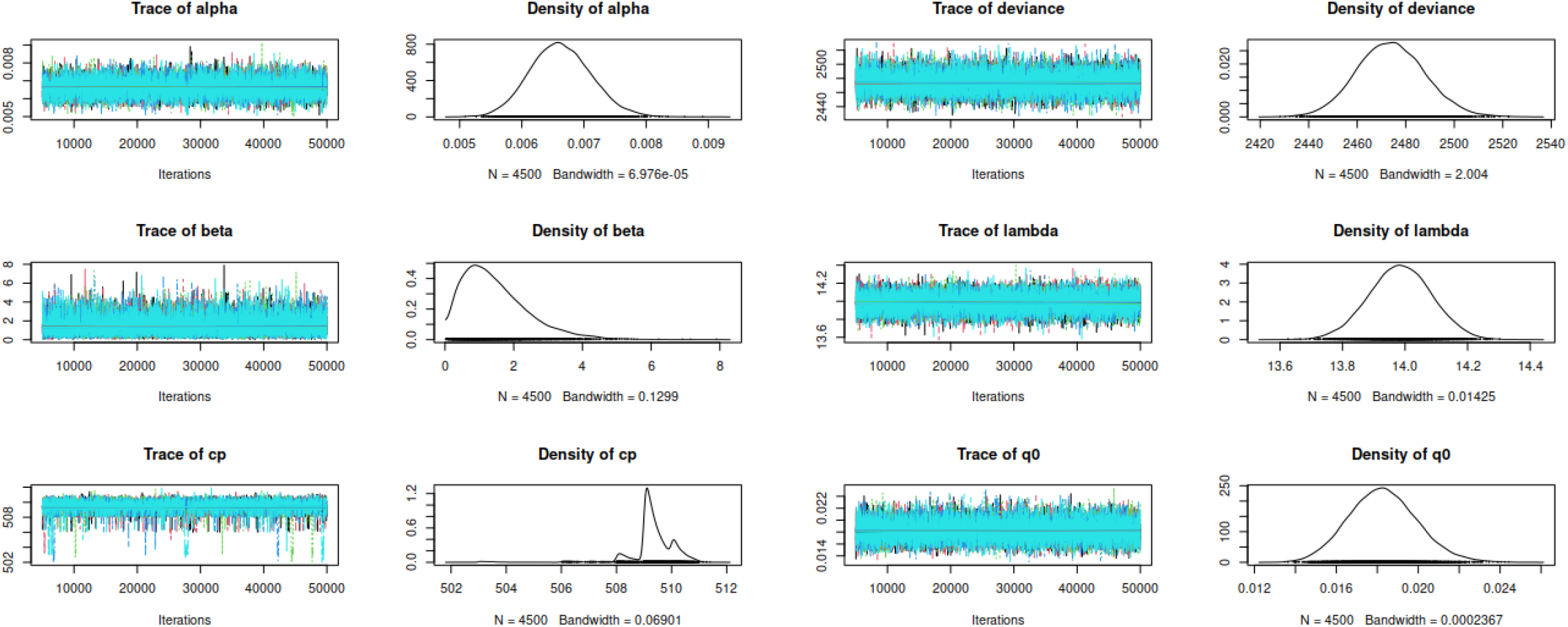
MCMC diagnostics: Chain convergence and posterior density functions for subarea F under scenario 1.

## 4 Discussion

Although various novel data science approaches have recently been proposed in seeking to obtain a better understanding of GBV, most focus primarily on identifying risk factors related to such episodes. To the best of our knowledge, this is the first time that advanced statistical modelling techniques have been used to estimate the actual magnitude of the scourge of GBV. The results of our study show that, on average, only around 11% of the victims of GBV between 2010-01-01 and 2021-12-31 in Barcelona’s *Àmbit Metropolità Nord* health area that requested primary healthcare attention were properly detected and registered (that is, 3,110 victims were diagnosed while the model estimates that there were a total of 27,264). The severity of this underreporting issue was especially dramatic at the beginning of this period –that is, before the professionals responsible for diagnosing such cases underwent awareness training– when only 3% of cases was registered. After the training provision, 42% of cases were registered. The impact of this awareness-raising activity can, therefore, be quantified as a 14-fold reduction in the underreporting issue. Although the importance of such training activities is manifest from the results of this and previous studies ([9, 10, 11, 12, 13]), the fact that its impact is best explained by the exponential shape of *q*_*t*_ seems to suggest that this training should be complemented with other activities focused on raising awareness of the actual magnitude of GBV, because otherwise it will take more than 20 years for the Catalan primary healthcare system to see the whole GBV iceberg. Another significant finding to emerge from the present study is the impact of mandatory confinement during the COVID-19 pandemic (March-June 2020) on episodes of GBV, estimated at an average growth of 1.25 cases per week (although subarea C might have experienced a growth rate of 3.9 cases per week, according to the corresponding 95% credible interval), a result that is in line with previous studies on the topic (see for instance [17]). Global concern has been expressed about this increase in GBV in relation to the mandatory home confinement imposed as a COVID-19 mitigation measure and a number of recommendations have been proposed aimed at controlling the incidence of GBV in future pandemics ([22]). Unfortunately, the results reported herein cannot be considered exclusive to the specific geographical area for which actual data were analyzed. On the contrary, these conclusions are straightforwardly generalizable to any other developed country of similar social characteristics ([7]), and may well be worse in developing countries ([23]). Whatever the case, the methodology proposed in this study can be used to analyze any given situation of interest, as all the code developed herein is freely available in the GitHub repository https://github.com/dmorinya/UnderreportedGBV.

## Data Availability

All generated code is available in a Github repository. The data used in this study is owned by the Catalan Institute of Health, and the authors are not authorized to distribute it. For any inquiries, please contact the corresponding author.

https://github.com/dmorinya/UnderreportedGBV

## Acknowledgements

This study has been funded by the Social Observatory of the “la Caixa” Foundation as part of the project LCF/PR/SR22/52570005. A.F-F acknowledges Agencia Estatal de Investigación for the financial support IJC2020-045188I/AEI/10.13039/501100011033 and a María Zambrano scholarship. This work was partially funded through the Severo Ochoa and María de Maeztu Program for Centers and Units of Excellence in R&D (CEX2020–001084-M) and by grant PID2022-137414OB-I00 fom the Spanish Government through the Ministerio de Ciencia e Innovación. The data were analyzed anonymously and all procedures were approved by the Ethics Committee of the IDIAP Jordi Gol research institute (23/198-P).

## 5 Supplementary Material

### 5.1 Model details

The number of cases diagnosed within the public primary healthcare system, *Y*_*t*_, is just a part of the actual process *X*_*t*_, expressed as

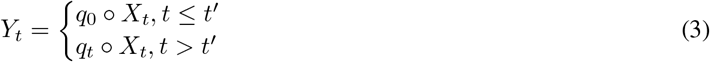

where ο is the *binomial thinning* operator, defined as 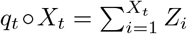, with *Z*_*i*_ independent and identically distributed Bernoulli random variables with probability of success *q*_*t*_and 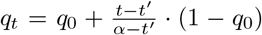 for *t > t*^*′*^, where *t*^*′*^ is the unknown change point at which the awareness training for primary healthcare professionals starts impacting the reported weekly number of diagnoses. Additionally, an alternative exponential modelling of *q*_*t*_ as 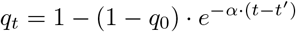 was considered and compared to the previous one in terms of the deviance information criterion (DIC).

### 5.2 Model diagnosis

The processes observed in each subarea under scenarios 2 and 3 are shown in Figures 11 and 12, respectively, together with a simulation based on the theoretical model described in the previous section and the parameter estimates for the corresponding scenario. It is evident that, as also occurred under scenario 1 (Figure 1), the theoretical model captures the behavior of the observed phenomenon accurately.

Model diagnostic and trace plots are shown in Figures 13 to 18. It can be seen that convergence is reached for all parameters and no particular patterns can be identified.

In addition, the potential scale reduction factor (PSRF) corresponding to all parameters in all subareas and considered scenarios are below 1.1, which is also indicative of the acceptable convergence of the MCMC chains. The PSRF is an estimated factor by which the scale of the current distribution for the target distribution might be reduced if the simulations were continued for an infinite number of iterations. Each PSRF declines to 1 as the number of iterations approaches infinity, and although the recommended proximity of each PSRF to 1 varies with each problem, the general goal is to achieve PSRFs below 1.1, as happens in this application.

### 5.3 Simulation study

To assess the performance of the model (3), a simulation study was conducted. The theoretical values for the parameter *q*_0_ ranged from 0.1 to 0.9; *α* = 1200, 1500, 2000 for linear *q*_*t*_ and *α* = 0.05, 0.1, 0.5 for exponential *q*_*t*_; *β* = 0.5, 2, 5; *λ* = 5, 7, 10; *t*^*′*^ = 100, 500, 900. For each combination of parameters, 100 random samples of size n=1000 were generated. The average relative bias, average interval length (AIL) and average 95% credible interval coverage are shown in Table 7. To provide a summary of the model’s robustness, these values were averaged over all combinations of parameters, considering their prior distribution to be a Dirac’s delta with all probability concentrated in the corresponding parameter value.

**Table 7:** Summary of model performance measures (average relative bias, average interval length (AIL) and average coverage) based on a simulation study for linear and exponential modelling of *q*_*t*_)

| $q_t$ | Parameter | Bias (%) | AIL | Coverage (%) |
| --- | --- | --- | --- | --- |
| Linear | $\alpha$ | 1.91 | 2207 | 90.1 |
| | $\beta$ | 0.002 | 3.98 | 96.4 |
| | $t'$ | 0.3 | 495 | 98.2 |
| | $\lambda$ | 0.0002 | 0.38 | 89.3 |
| | $q_0$ | 0.00004 | 0.07 | 89.1 |
| Exponential | $\alpha$ | 0.0002 | 0.93 | 99.4 |
| | $\beta$ | 0.002 | 4.19 | 97.4 |
| | $t'$ | 0.089 | 155 | 98.0 |
| | $\lambda$ | 0.0002 | 0.35 | 95.5 |
| | $q_0$ | < 0.0001 | 0.083 | 93.6 |

As shown in Table 7, the model behaves as expected regardless of whether *q*_*t*_ is modelled linearly or exponentially. In general, the 95% credible intervals coverage is high, even with relatively narrow intervals and low relative bias.

